# Prevalence of SARS CoV-2 infection among Health Care Workers of a hybrid tertiary COVID 19 hospital in Kerala

**DOI:** 10.1101/2021.07.19.21260792

**Authors:** S J Jessy, M Shamha Beegum, S Genga, G Bindu, S Chintha, Sukshma Sasidharan, Ansu Tonio, R Aravind

## Abstract

**Back ground and objectives:** This study was undertaken to estimate the prevalence of SARS-CoV-2 infection among Health care workers [HCWs] of a hybrid COVID treatment hospital in Kerala.

**Methods:** The study was conducted during 3^rd^ week of January 2021. Among 3550 HCWs, 979 subjects were selected by stratified random sampling and grouped into high risk and low risk category based on job setting. Demographic details and clinical information regarding previous history of COVID 19 were collected at the time of SARS-CoV-2 IgG testing.

**Results:** From 979 subjects, the data with respect to 940 health care workers were analysed. SARS-CoV-2 IgG was detected in 19.1% of HCWs. Seroprevalence among high risk group was 20.3% and that in low risk group was 7.4% [p=0.005]. In high-risk group, seropositivity was noted in 30.54 % of nurses, 19% hospital attenders, 18.9% resident doctors and 6.4% consultant doctors. In those with past history of SARS-CoV-2 infection, seropositivity was 75.4%. In those who were COVID positive during July2020, 33.3% were still IgG reactive.

**Interpretation and conclusion:** The study reported 19.1% SARS CoV-2 IgG reactivity among health care workers in our hospital. Seropositivity was significantly higher in high risk group compared to low risk group. Antibody decay kinetics in our study is comparable to that in published literature. Infection control challenges in hybrid hospitals account for higher seropositivity in this study compared to overall seroprevalence among HCWs in Kerala.

## Introduction

COVID 19 is a multisystem disease with predominant involvement of the respiratory system. COVID 19 transmission dynamics include droplet, contact and airborne transmission in specific settings. Health care workers are in close contact with COVID 19 patients, and so the probability of getting infected is very high as evidenced by studies from across the world. As around 40% of SARS-COV2 infections are asymptomatic, the exact prevalence among HCWs can be identified only by a seroprevalence study.

The first national population based serosurveillance study conducted by ICMR during May-June 2020, found that 0.73% of adults in India were exposed to SARS -COV2 infection [1]. SARS-COV 2 IgG seroprevalence study conducted by ICMR across three districts of Kerala in May, August and December 2020 showed prevalence of 0.33%, 0.73% and 11.6% respectively. The same survey showed that the seroprevalence in India during same time period was 0.8%, 6.60% and 21.50%. Majority of the studies from across the world have shown that seroprevalence among HCWs is higher than that in the community. This is because Health care workers (HCW] have more risk of exposure, and those working in critical care units are likely to have exposure to higher viral inoculum load from aerosol generating procedures which are inevitable. Risk of acqusition of HCW infection is higher in those who work in hybrid hospitals compared to that in designated COVID hospitals. This is because in hybrid hospitals both COVID and non-COVID cases are admitted where as in designated COVID hospitals only confirmed COVID 19 cases are admitted. Infection prevention and control is more challenging in hybrid hospitals than in designated COVID hospitals.

Seroprevalence among HCWs vary from hospital to hospital and depends on the stage of the pandemic in the district during the study period and infection control practices adopted in the hospital. In India, a study done among 25% of HCW of a tertiary care hospital had shown seroprevalence of 11.94% [2]. According to ICMR third round seroprevalence study, 25.7% of HCWs in India have COVID 19 antibodies. A study conducted during July 2020 among HCWs of a hospital in Kerala showed no prevalence of SARS COV-2 IgG [3]. But during the study period seroprevalence in Kerala was only 0.2%. There is no published study related to seroprevalence of COVID 19 among HCWs from a hybrid COVID hospital in Kerala after surge of cases occurred and population seroprevalence increased from 0.2 to 11.6%. This study was undertaken to estimate the prevalence of SARS-CoV-2 infection among HCWs of Government Medical College Thiruvananthapuram [GMCT], a hybrid tertiary care centre. Thiruvananthapuram district has reported maximum number of cases of COVID 19 in Kerala and had peak case load in October 2020. Eventhough the protective role of antibodies against SARs-CoV-2 is unknown; they can be taken as a correlate of antiviral immunity and may correspond to plasma viral neutralizing activity. Persistence and decay kinetics of SARS-CoV-2 IgG among HCWs needs to be studied in detail.

## Materials and Methods

**Study design**: cross sectional study

**Study setting**: Central Biochemistry Lab GMCT.

**Study Period**; Jan 8, 2021 to Jan 19, 2021

**Study population** – HCWs at GMCT

**Sample size**

Sample size was calculated using equation

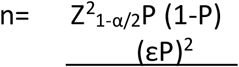

Where P= anticipated population proportion

ε= relative precision (it is recommended to fix the value εfrom a minimum of 10% to a maximum of 20%.

|  |  |
| --- | --- |
| Expected Proportion | 0.11 |
| Relative Precision (%) | 20 |
| Desired confidence level (1- alpha) % | 95 |
| Required sample size | 979 |

Expected prevalence-11%

[Based on study done by Khan MS, Haq I, Qurieshi MA, Majid S, Bhat AA, Obaid M, et al. SARS-CoV-2 seroprevalence in healthcare workers of dedicated-COVID hospitals and non–COVID hospitals of District Srinagar, Kashmir]

### Sampling

From 3550 HCWs at GMCT, 979 HCW were selected by stratified random sampling. Risk categorization into high-risk and low risk was done based on job setting. High risk group HCWs was defined as those involved in direct care of patients with COVID 19 and low risk group included HCWs working in non-COVID pool. HCWs who were not willing to participate were excluded from the study. After obtaining informed written consent, data with regard to demographic variables, clinical history with regard to symptomatology in case of past history of COVID 19, infection prevention and control measures adopted etc were captured on to a structured proforma approved by Institutional Ethics Committee. Anti-SARS-CoV-2 IgG test was performed on serum by chemiluminescent immune assay [CLIA] using VITROS Anti-SARS-CoV-2 IgG reagent pack and VITROS Anti-SARS-CoV-2-IgG calibrator on VITROS ECi 3600 Immunodiagnostic system. Quality checks and calibration were done as per CLSI guidelines. VITROS Anti-SARS CoV-2 IgG Assay kit used for the study has US FDA EUA, CE certification and is recommended by ICMR for Sero-surveillance purpose. The sensitivity and clinical specificity of the assay kit is 90% and 100% respectively. Using VITROS immunodiagnostic system, results are calculated automatically as signal/Cut off (S/Co values). In a serosurvey study on convalescent plasma therapy done for COVID19 at Mayo clinic using the VITROS IgG kit, a S/Co value between 1.0-4.64 denoted low tite, 4.62-18.45 medium titer and above 18.45 high titer.

### Data analysis

Statistical analysis: - Numerical variables such as age, IgG values etc were presented as mean and standard deviation. Categorical variables such as sex, risk category of subjects, symptoms, co-morbidities and test results were presented as frequency and percentage. 95% CI of observed prevalence and true prevalence was adjusted for sensitivity and specificity of the reagent kit. The corresponding 95 % CI for adjusted prevalence was calculated by Blaker’s method using R software. IgG reactivity among various categories of participants were analyzed by Chi square test. A two-sided probability value of <0.05 was considered statistically significant. Random sample numbers were generated and data analysis was performed Using R software (R version 3.6.2 (2019-12-12) Copyright (C) 2019 The R Foundation for Statistical Computing Platform: x86_64-w64-mingw32/x64 (64-bit)) and SPSS version 16.0.

## RESULTS

The sample size calculated for the study was 979. Of this, 39 participants were excluded from the study due to incomplete proforma and so the final sample size was fixed at 940.

**Figure 1.**
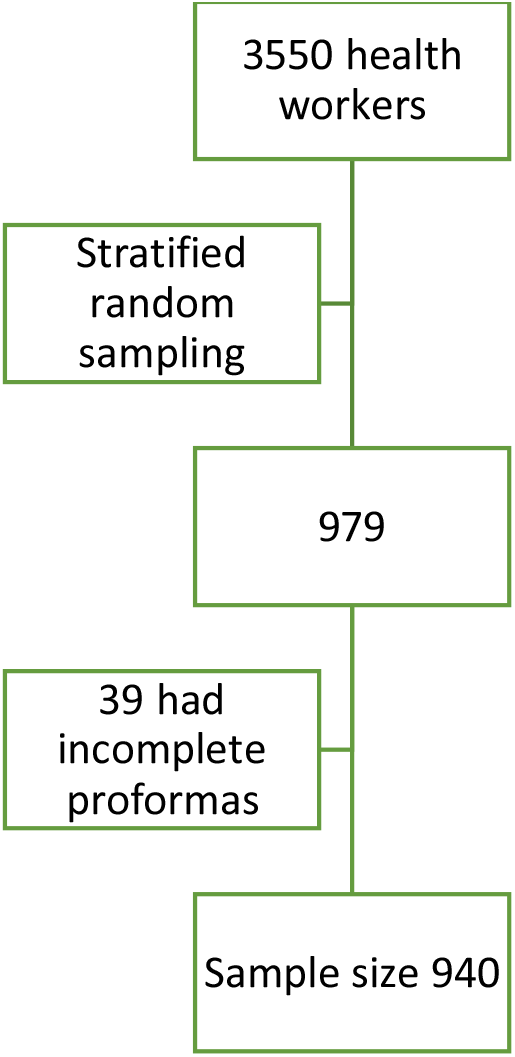
Algorithm showing sample size taken for statistical analysis

In the study SARS-CoV-2 IgG seropositivity was identified in 180 cases, resulting in an observed prevalence of 19.1%. Reagent sensitivity and specificity was 90% and 100% respectively. When 95% CI is adjusted for test sensitivity and specificity by Blaker’
ss method, the true prevalence arrived at is 21.2%.

Out of the 940 study participants (Table 1), 765 (81.4%) were below 50 years and 175(18.6%) were above 50 years of age. Sex ratio among study participants was 69.3% females and 30.6% males respectively.

**Table I.** Baseline characteristics

| N=940 |  |  |
| --- | --- | --- |
|  |  | n(%) |
| Age in years | ≤50 | 765(81.4) |
|  | >50 | 175(18.6) |
| Sex | Male | 288(30.6) |
|  | Female | 652(69.4) |
| Study participants |  |  |
| High risk group | Doctors at high risk | 140(14.9) |
|  | Residents | 312(33.2) |
|  | Nurses | 249(26.5) |
|  | Attenders | 158(16.8) |
| Low risk Group | Doctors at low risk | 26(2.8) |
|  | Technicians | 39(4.1) |
|  | JLAs | 16(1.7) |
| Category | High risk | 859(91.4) |
|  | Low risk | 81(8.6) |

Study participants were categorized into high-risk group (n= 859, 91.3%) and low risk group (n= 81, 8.6%).

Among the high-risk category, 140 (14.89%) were doctors, 312 (33.2%) were residents, 249 (26.4%) were nurses and 158(16.8%) were hospital attenders. Low risk category included doctors (n= 26, 2.8%), technicians (n=39, 4.4%) and junior lab assistants (n= 16, 1.6%)

*Past history of COVID test positivity and seropositivity*-(Table 2).In the study, 134 participants had been tested positive by RTPCR in the past for SARS-CoV -2 infection. Among these 134 study subjects, 101 (75.4%) were SARS-CoV-2 IgG reactive. Of the 536 subjects who had tested COVID negative in the past, 51 (9.5%) were seropositive for SARS-CoV -2 IgG. In 270 subjects who had never undergone COVID test in the past, 28 (10.4%) were seropositive (p< 0.001}

**Table 2.**
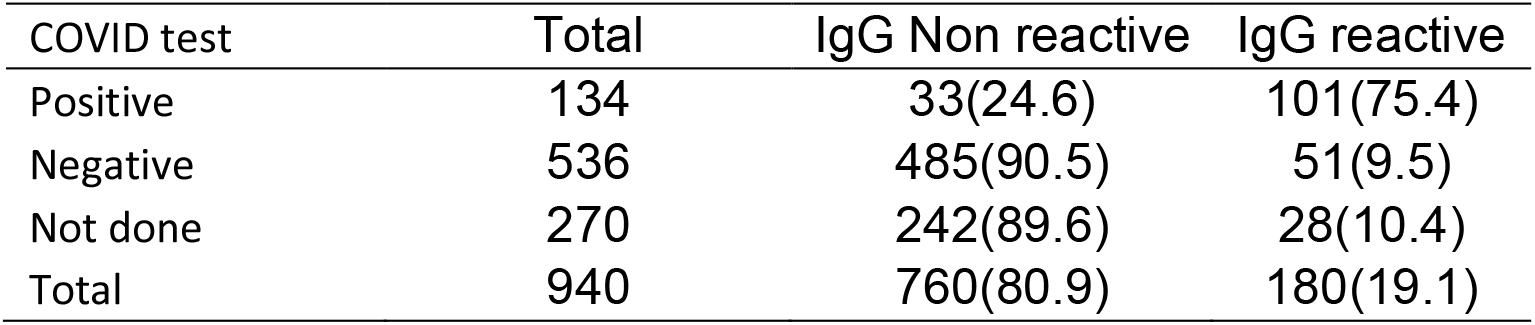
Association of COVID IgG positivity and COVID 19 test

*Prevalence as per age& sex*- (Table 3) Seropositivity below 50 years was 19.3% (n=148) and above 50 years was 18.3 % (n= 32) with a p- value of 0.748 which is not statistically significant. Seropositivity among males and females were 14.9% and 21% respectively with a p value of 0.029 which is statistically significant.

**Table 3.**
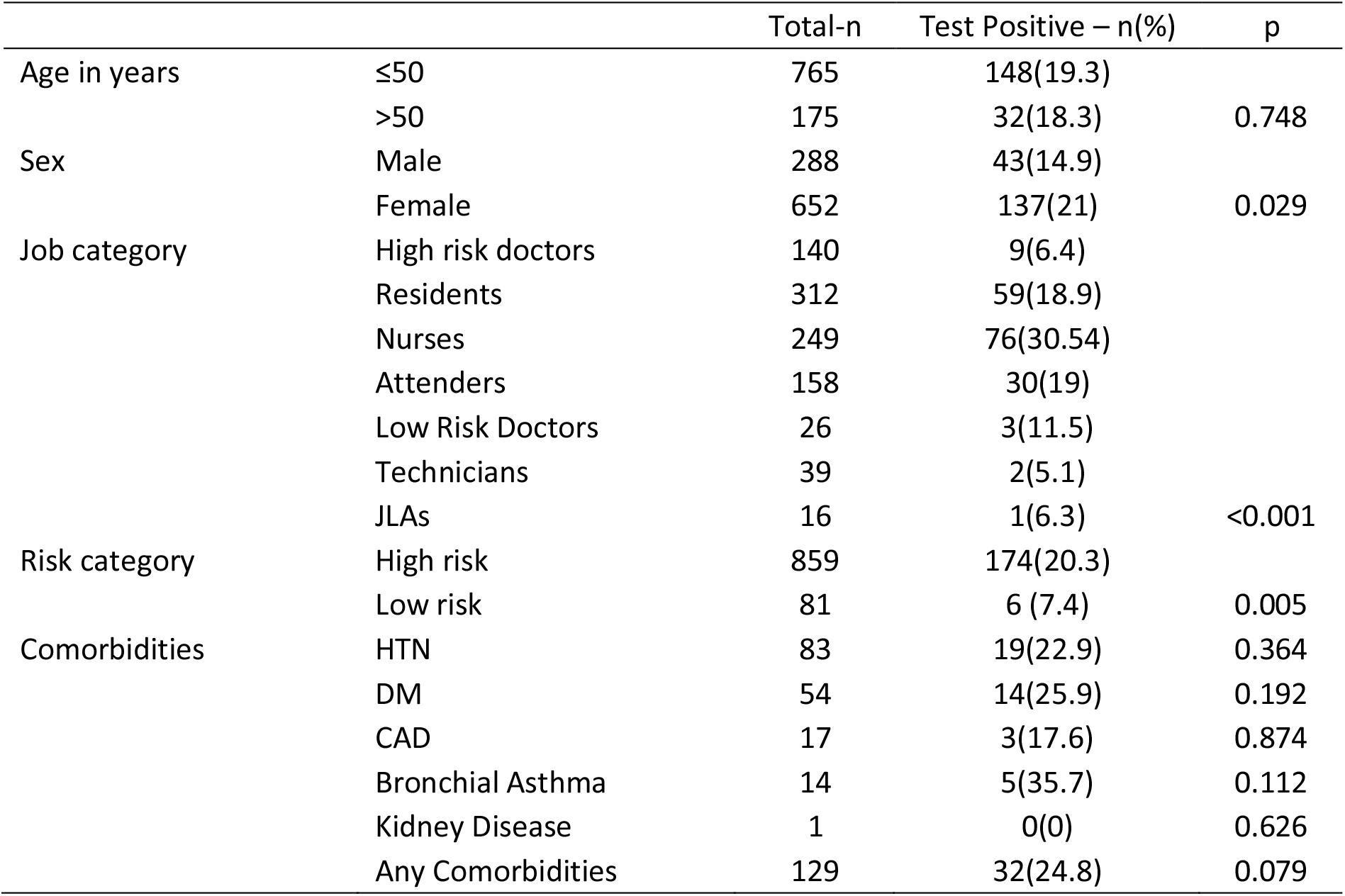
Factors associated with SARS-CoV-2 IgG positivity

*Seropositivity based on risk and job category*-(Table 3)

Among HCWs in High- risk group 20.3% (n= 174) and among low risk group 7.4% (n=6) were seropositive with a stastically significant p value of 0. 005. Among high-risk group, 6.4% of Doctors (n= 9), 18.9% residents (n=59), 30.54% of nurses(n=76) and 19% of attenders (n=30) were seropositive with a stastically significant p value <0.001.

In the low risk group, 11.5% of doctors, 5.1% of technicians and 6.3% of junior lab assistants were seropositive.

*Seropositivity and comorbidities* – (Table 3) Among 180 seropositive subjects, 24.8% had atleast one comorbidity. Of them 22.9% were hypertensives, 25.9% were diabetic, 17.6% had coronary artery disease and 35.7% had bronchial asthma.

*Symptomatology in SARS-COV 2 IgG seropositive subjects* - (Table 4)

**Table 4.** Association of COVID 19 positivity, COVID 19 symptoms and symptoms among who were COVID test positive

|  |  | Total-n | Test Positive – n(%) | p |
| --- | --- | --- | --- | --- |
| COVID 19 Symptoms | Fatigue | 123 | 51(41.5) | <0.001 |
|  | Sore throat | 170 | 53(31.2) | <0.001 |
|  | Running Nose | 141 | 44(31.2) | <0.001 |
|  | Cough | 105 | 36(34.3) | <0.001 |
|  | Fever | 160 | 72(45) | <0.001 |
|  | Anosmia | 46 | 32(69.6) | <0.001 |
|  | Ageusia | 23 | 18(78.3) | <0.001 |
|  | Vomiting | 10 | 6(60) | 0.001 |
|  | Diarrhea | 35 | 9(25.7) | 0.314 |
|  | Any Symptoms | 353 | 115(32.6) | <0.001 |
| Symptoms among COVID test positives | Fatigue | 48 | 38(79.2) | 0.446 |
|  | Sore throat | 46 | 40(87) | 0.024 |
|  | Running Nose | 30 | 24(80) | 0.504 |
|  | Cough | 25 | 22(88) | 0.104 |
|  | Fever | 59 | 48(81.4) | 0.154 |
|  | Anosmia | 36 | 31(86.1) | 0.08 |
|  | Ageusia | 20 | 17(85) | 0.279 |
|  | Vomiting | 6 | 5(83.3) | 0.643 |
|  | Diarrhea | 11 | 9(81.8) | 0.605 |
|  | Any Symptoms | 93 | 74(79.6) | 0.089 |
|  | No symptoms | 41 | 27(65.9) | 0.089 |

Of the subjects who were seropositive for SARS-COV-2 IgG, 41.5% had fatigue, 31.2% sore throat, 31.2% rhinorrhoea, 34.3% cough,72% fever, 69.6% anosmia, 78.3% ageusia, 60% vomiting, 25.7% diarrhea and 32.6% had atleast one of the above symptoms during the last 6 months.

*Symptomatology in subjects who had tested positive for COVID 19 and are seroreactive* - In COVID tested positive subjects who were seroreactive 88% had cough, 87% sore throat, 86.1% anosmia, 85% ageusia, 81.4% fever, rhinorrhoea 80%, and 79.2% had fever at the time of diagnosis of COVID 19.

*Trends in SARS-CoV-2 IgG antibody reactivity in relation to month in which subjects were detected to have COVID 19* (Table 5)

**Table 5.**
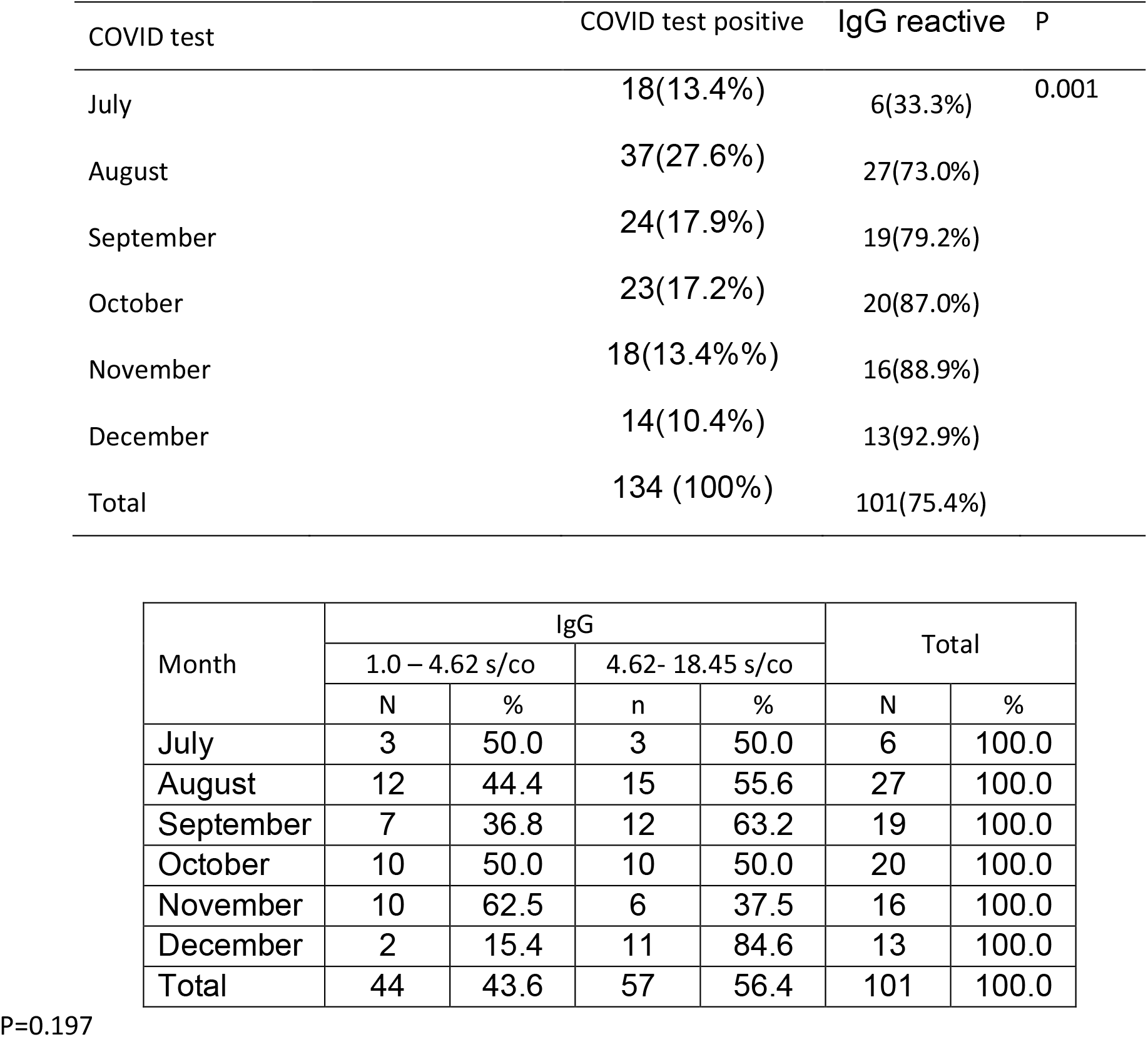
Trend in COVID IgG antibody reactivity among cases detected from July to December 2020

| COVID test | COVID test positive | IgG reactive | P |
| --- | --- | --- | --- |
| July | 18(13.4%) | 6(33.3%) | 0.001 |
| August | 37(27.6%) | 27(73.0%) |  |
| September | 24(17.9%) | 19(79.2%) |  |
| October | 23(17.2%) | 20(87.0%) |  |
| November | 18(13.4%) | 16(88.9%) |  |
| December | 14(10.4%) | 13(92.9%) |  |
| Total | 134 (100%) | 101(75.4%) |  |

| Month | IgG |  |  |  | Total |  |
| --- | --- | --- | --- | --- | --- | --- |
|  | 1.0 – 4.62 s/co |  | 4.62- 18.45 s/co |  |  |  |
|  | N | % | n | % | N | % |
| July | 3 | 50.0 | 3 | 50.0 | 6 | 100.0 |
| August | 12 | 44.4 | 15 | 55.6 | 27 | 100.0 |
| September | 7 | 36.8 | 12 | 63.2 | 19 | 100.0 |
| October | 10 | 50.0 | 10 | 50.0 | 20 | 100.0 |
| November | 10 | 62.5 | 6 | 37.5 | 16 | 100.0 |
| December | 2 | 15.4 | 11 | 84.6 | 13 | 100.0 |
| Total | 44 | 43.6 | 57 | 56.4 | 101 | 100.0 |

P=0.197

Of the 134 subjects who were diagnosed to have COVID 19 in the past, 101 [75.4%] were detected to be seropositive for SARS-CoV-2 IgG during the study period [8-1-2021 to 19-1-2021]. Seropositivity among subjects detected to have COVID 19 in July, August, September, October, November and December were 33.3%, 73%,79.2%,87%,88.9% and 92.9% respectively. A low titre S/Co was present in 43.6% and medium titre S/Co was present in 56.4% of study subjects with past history of COVID 19 who were SARS-CoV-2 IgG reactive. None of the study subjects had a high titre S/Co.

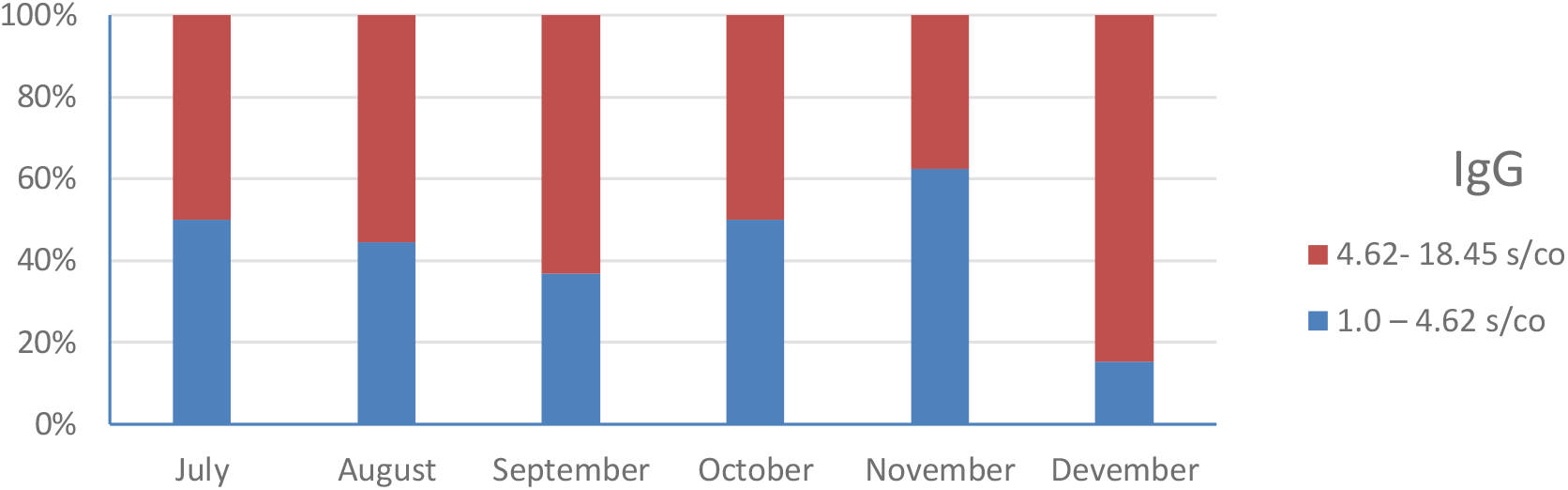

*S/Co values of SARS-CoV-2 IgG reactive HCW* (Table 6)

**Table 6.**
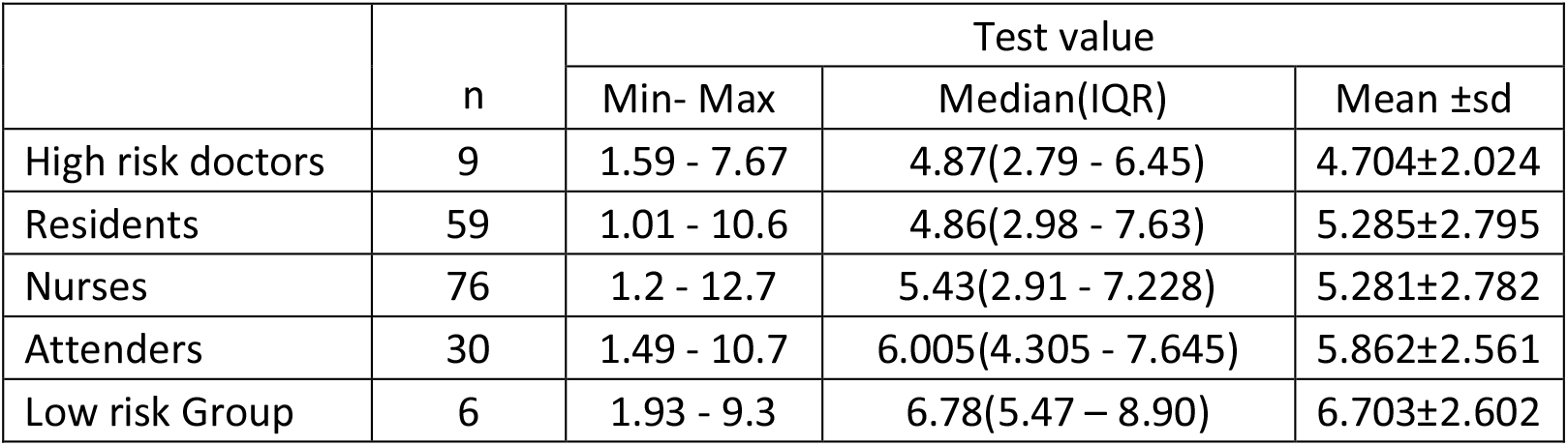
COVID IgG Antibody titer across risk categories.

|  | n | Test value |  |  |
| --- | --- | --- | --- | --- |
| | | Min- Max | Median(IQR) | Mean ±sd |
| High risk doctors | 9 | 1.59 - 7.67 | 4.87(2.79 - 6.45) | 4.704±2.024 |
| Residents | 59 | 1.01 - 10.6 | 4.86(2.98 - 7.63) | 5.285±2.795 |
| Nurses | 76 | 1.2 - 12.7 | 5.43(2.91 - 7.228) | 5.281±2.782 |
| Attenders | 30 | 1.49 - 10.7 | 6.005(4.305 - 7.645) | 5.862±2.561 |
| Low risk Group | 6 | 1.93 - 9.3 | 6.78(5.47 – 8.90) | 6.703±2.602 |

Among *SARS-CoV-2 IgG reactive HCWs*, 40.6% had value between 1.0-4.62 [low titre] and 59.4% had value between 4.62-18.45 [medium titre]. No subjects had high S/Co titre in this study.

The relation between time since infection and antibody titre is shown in figure 2. Antibody titre of subjects infected 6 months prior to testing showed 33% positivity with rates inversely proportionate to time from infection. At 1 month after infection, only one patient had negative titre.

**Figure 1.**
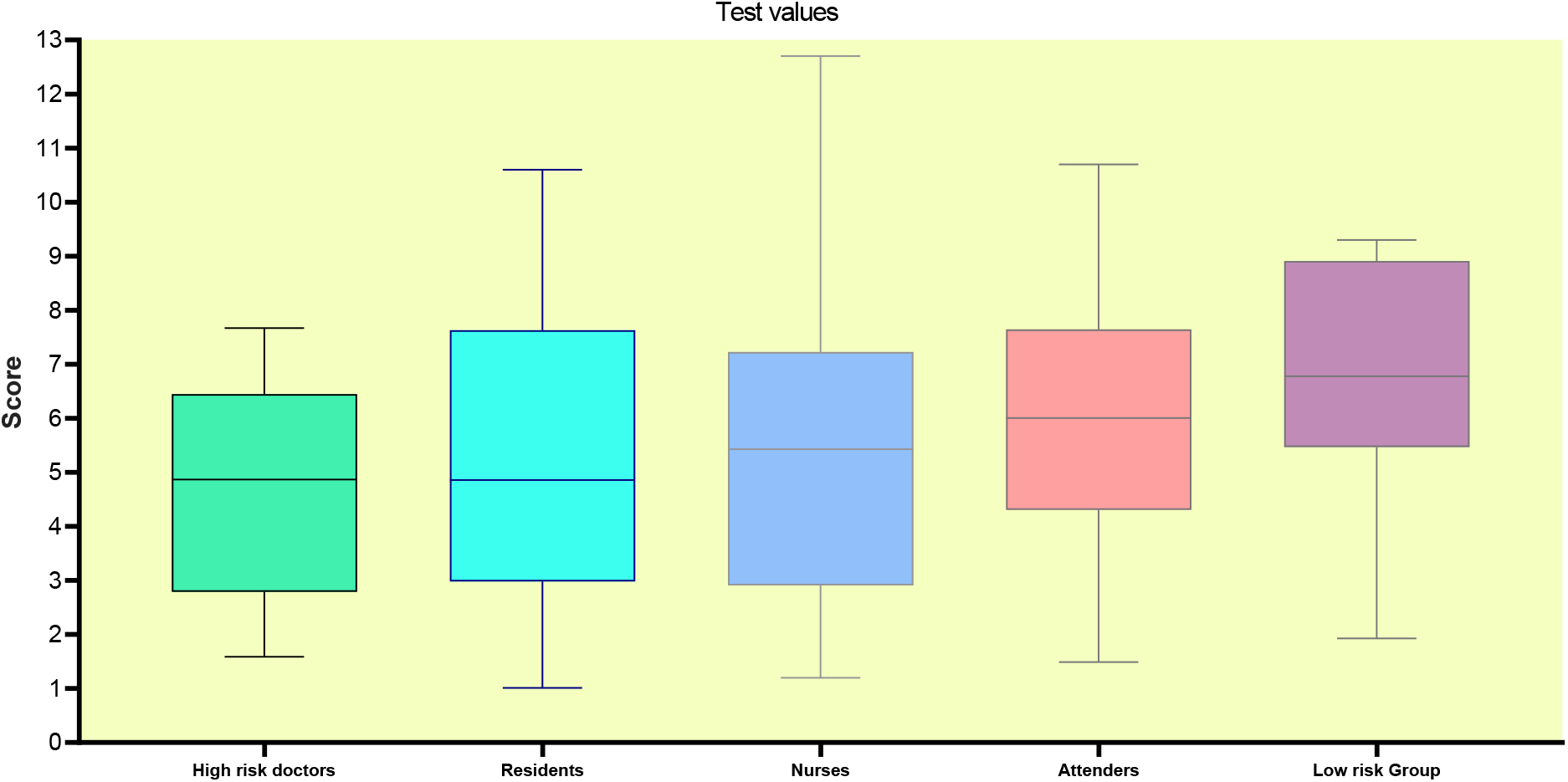
Box plot representing SARS-CoV-2 IgG Antibody titres across risk categories.

**Figure 2.**
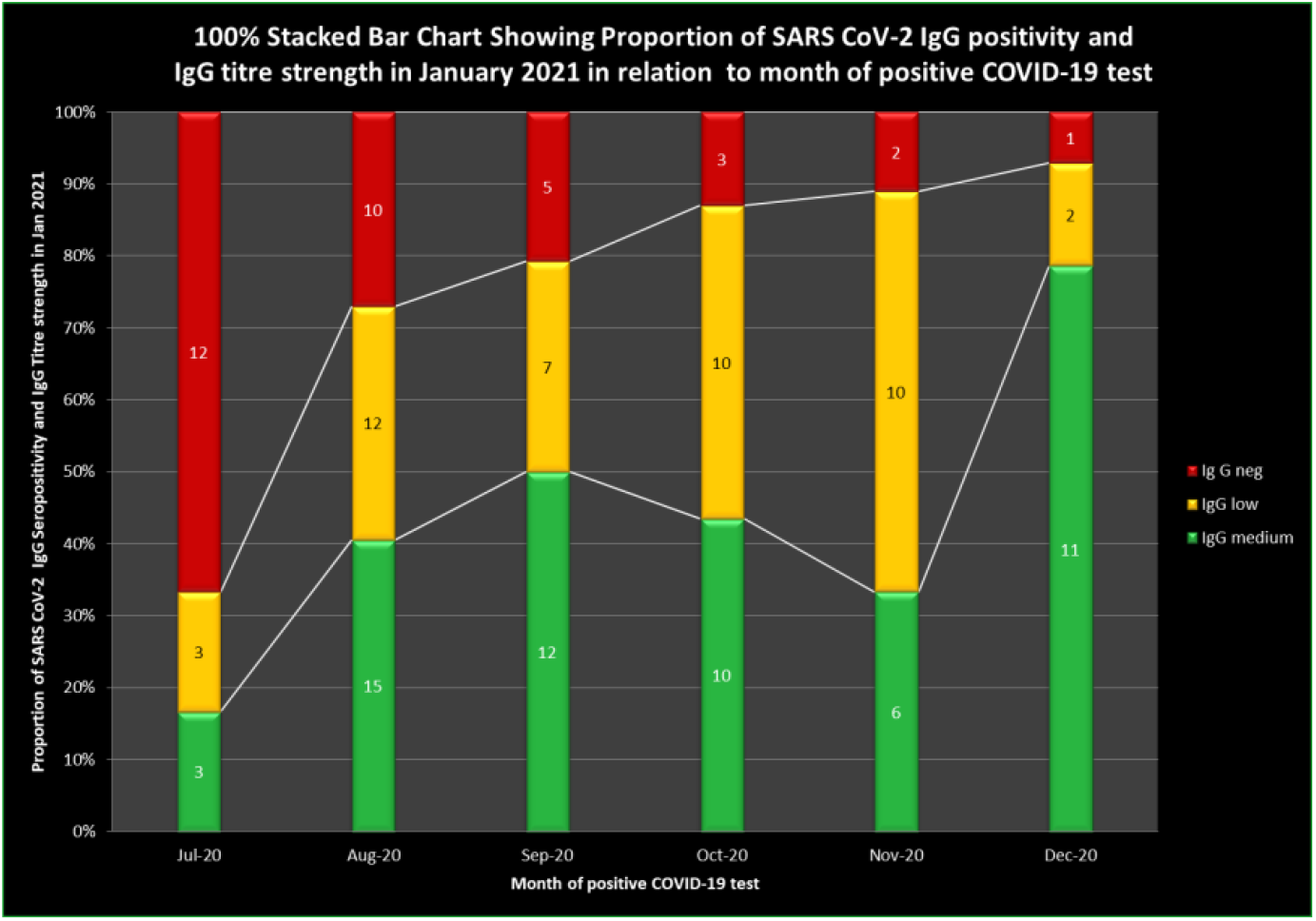
100% stacked bar chart showing the proportion of SARS CoV 2 IgG positivity and IgG titre strength in January 2021 in relation to month of positive COVID 19 test.

When COVID Anitibody positivity titres were analyzed among symptomatic and asymptomatic groups, antibody titres showed a decrease with time from infection in both groups and are depicted in Figure 3, 4 and 5.

**Figure 3.**
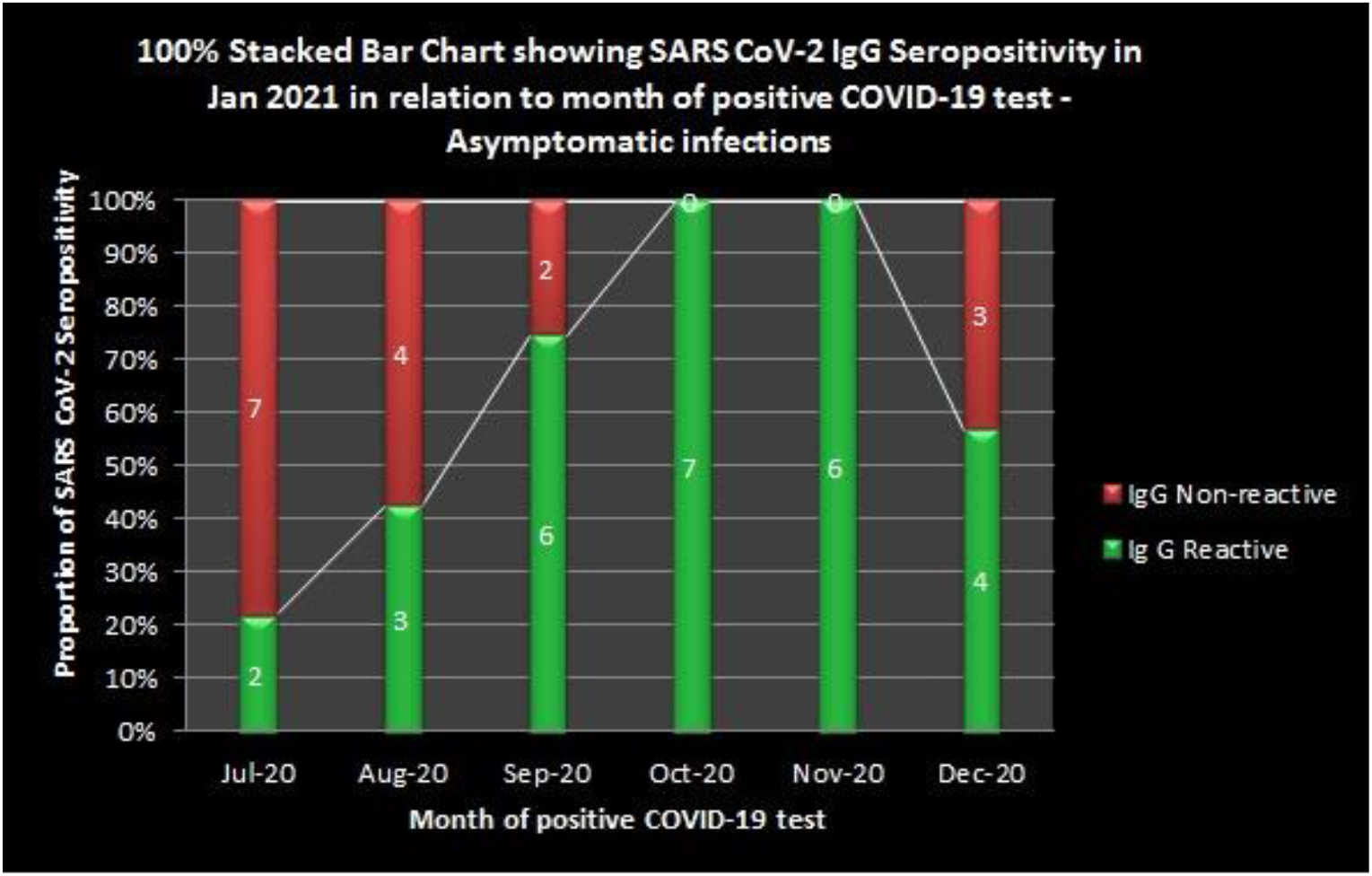
100% stacked bar graph showing SARS CoV 2 IgG sero positivity in Jan 2021 in relation to month of positive COVID 19 test – Asymptomatic infection.

**Figure 4.**
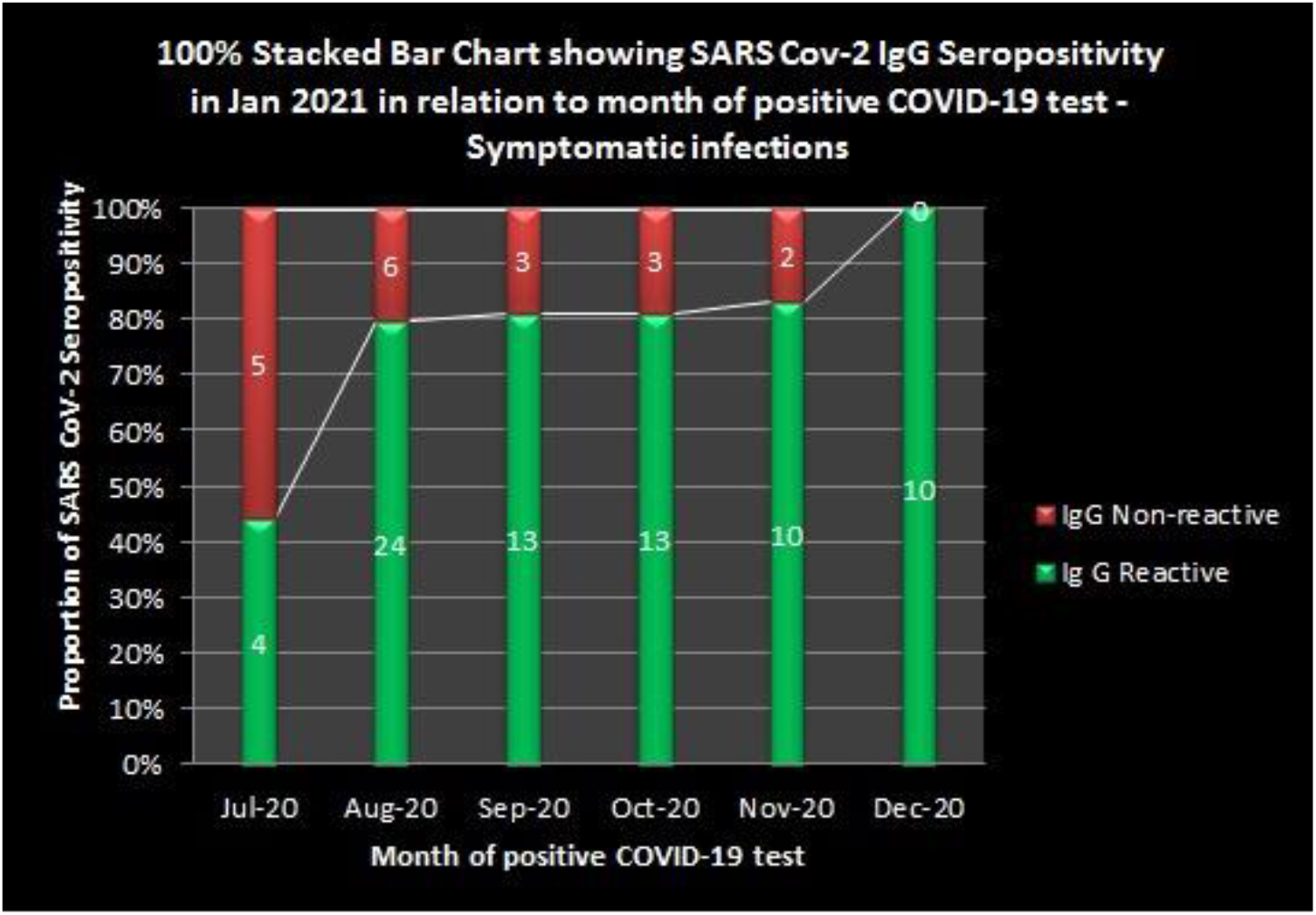
100% stacked bar graph showing SARS CoV 2 IgG sero positivity in Jan 2021 in relation to month of positive COVID 19 test – Symptomatic infection.

**Figure 5.**
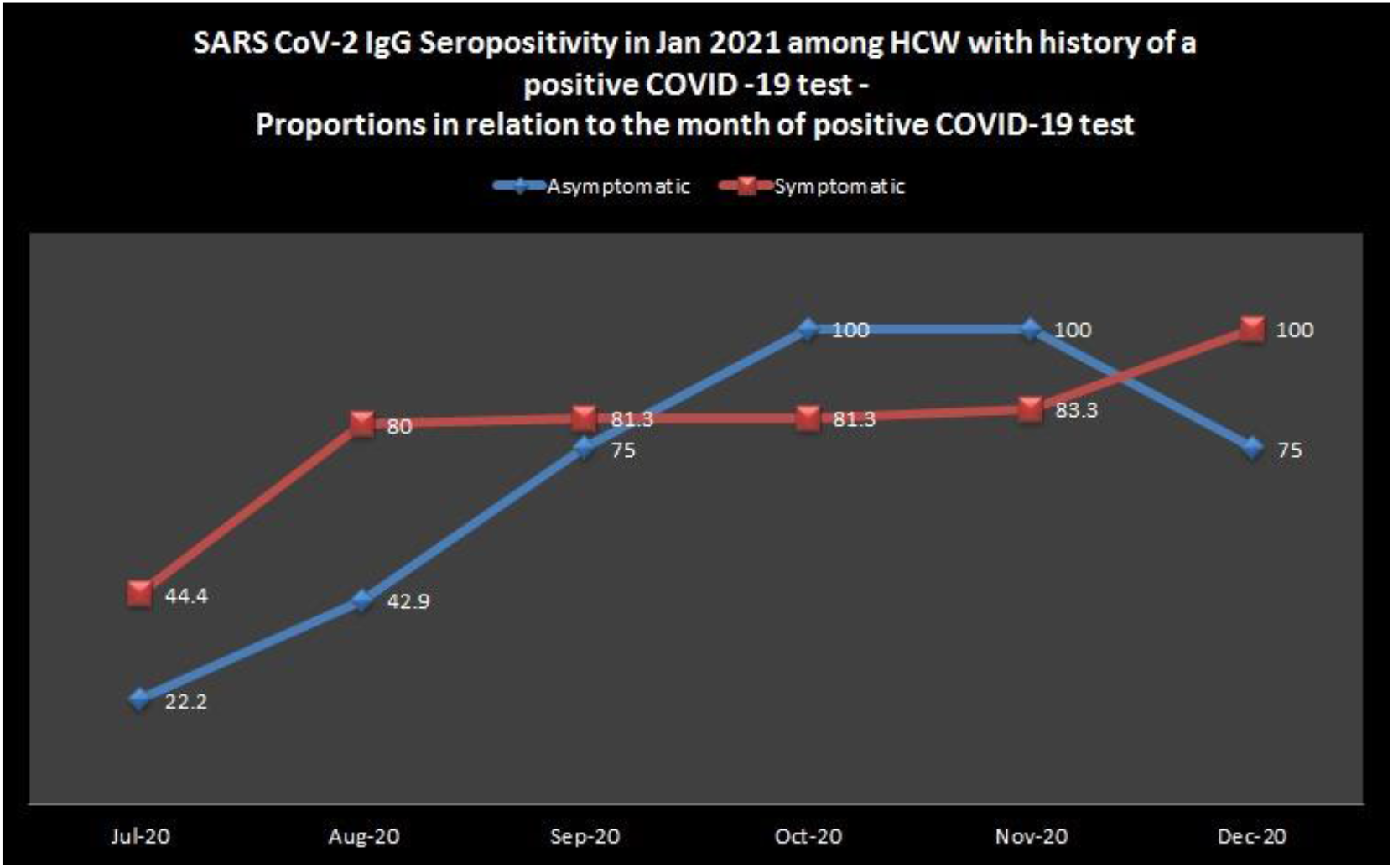
Showing SARS CoV 2 IgG seropositivity in Jan 2021 among HCW with history of a positive COVID 19 test – proportions in relation to the month of positive COVID 19 test.

## Discussion

Kerala was the first state in India to report COVID 19 case on January 29, 2020. First COVID 19 patient got admitted to GMCT on March 13^th^. GMCT, a 3250 bedded tertiary care hospital, was converted to a hybrid COVID hospital catering to both non-COVID as well as patients with moderate and severe SARS-CoV 2 infection. Being a border district and due to multiple coastal clusters, Thiruvananthapuram district had the maximum number of cases in Kerala with epidemic peak in October, 2020. Till date more than 10000 COVID 19 patients have been admitted to GMCT and 936 health care workers have been diagnosed to have COVID 19. In a study done during May 2020 in Kerala, the prevalence of SARS-CoV 2 infection was only 0.5% among HCWs (technical paper, Kerala). At the time of the study, total number of cases in Kerala was around 1000 only. Seroprevalence of SARS-CoV 2 infection in Kerala which was 0.3% in August increased to 11.6% in December and during this period, a similar increase in COVID 19 among HCWs was noted.

The present study was conducted between 8^th^January and 19^th^ January 2021.As on 19^th^ January, Kerala had a cumulative case load of 8,57,380 cases with 70,259 active cases. Our study showed a seropositivity of 19.1% among HCWs. A similar study done at National capital region of India conducted between January 12^th^ and February 13^th^ 2021 reported a seroprevalence of 46.2% among HCWs [4]. In India, the seropositivity among HCWs was 2.5% during June in Kashmir, 11% in Mumbai during May and 11.94% in Kolkata during July 2020 [5,6,3]. Lower seroprevalence among HCWs in these studies reflect the stage of the pandemic in respective states during the study period. The seropositivity among HCWs in other countries was 1.6%, 3.4% and 6% in Germany, Italy and England respectively [7, 8, 9].This low prevalence of SARS-CoV-2 IgG antibodies reflects the burden of pandemic in the study population in the particular time window. In Kerala, COVID 19 peaked in October 2020 and the epidemic curve has plateaued ever since. Higher SARS-CoV -2 IgG seropositivity among HCWs observed in our study reflects the epidemic situation in the state in the months prior to study period and that is the reason why seropositivity among HCWs in our study is high. Interestingly in the seroprevalence study conducted by Government of Kerala in February 2021, seropositivity among HCWs was 10.5%.The high seropositivity among HCWs [19.1%] in GMCT compared to the average seropositivity among HCWs [10.5%] across the state reflects the infection prevention and control challenges in a hybrid hospital which caters to both COVID and non-COVID patients.

In this study statistically significant difference in seropositivity was observed among female HCWs [21%] compared to that in males [14.9%]. This is due to the greater representation of female nurses in high risk category subgroup who turned seropositive [30.54%] .Initial studies from China had reported a sex disparity in COVID 19 epidemiology, whereas global Health 50/50 research initiative has observed similar number of cases in men and women [10].

In our study, statistically significant difference in seropositivity [20.3%] is observed in high risk category compared to that in low risk category [7.4%]. This finding is consistent with a study done in Switzerland [11]. Among the subgroups of high risk category, seropositivity was higher in nurses [30.54%] followed by hospital attenders [19%], resident doctors [18.9%] and consultant doctors [6.4%]. Nurses are at the highest risk of occupational exposure due to the nature of their work which results in longer duration and intensity of exposure to SARS-CoV-2.This is the reason why among HCWs in high risk category, nurses have maximum seropositivity. Consultant doctors have minimum duration of exposure to COVID 19 patients and that explains why seropositivity among them is only 6.4%.

In our study of the 180 subjects who were seropositive, only 24.8% had atleast one co-morbidity. This can be explained by the fact that 81.4% of study population was below 50 yrs of age. More over in our hospital, younger staff without co-morbidities were posted preferentially in high-risk areas. The practice of selectively posting those with co-morbidities in low risk areas in our institution probably helped in protecting them from getting infected.

The potential symptoms suggestive of COVID 19 in seropositive HCWs who had never tested for COVID 19 were analyzed in this study. Ageusia and anosmia were the common symptoms in that group. The predictive symptomatology for COVID 19in this study is similar to that in studies done in Sweden [12] and a multinational population-based cohort in US & UK [13]. In this study, in seropositive with past history of COVID 19 all the potential COVID 19 symptoms were present in more than 75% of subjects similar to a study done on health care personnel in Washington [14].

In this study, of the 134 HCWs with past history of COVID 19, 75.4% were detected to be seropositive. SARS-CoV-2 IgG was tested by CLIA method. Different methods to measure SARS-CoV-2 IgG like ELISA, CLIA and rapid lateral flow immunoassay have been found to have equivalent clinical performance for detecting IgG 14 days after onset of symptom in certain studies. [15]. Whereas a systematic review and meta-analysis to determine diagnostic accuracy of serological tests for COVID 19 detection have found lower sensitivity for lateral flow immunoassay when compared to ELISA and CLIA [16].

In this study, 75.4% of the subjects with past history of COVID -19 were seropositive for SARS-CoV-2 IgG. Antibody decay kinetics in our study assuming that all patients with past history of COVID 19 had developed IgG antibodies, is comparable to the rate of decay in published literature [17]. Even after six months of SARS-CoV-2 infection, 33.3% of patients had SARS-CoV-2 IgG antibody. The persistence of SARS-Cov-2 IgG in patients who were diagnosed to have COVID 19 at 5,4,3,2 and 1 month prior to the study period was 73%, 79.2%, 87%, 88.9% and 92.9% respectively. Initial studies on antibody decay kinetics in COVID 19 had shown that majority of the patients seroreverted by three months [17]. Similar antibody kinetics was observed in another study performed on 271 lab confirmed SARS-CoV-2 infection [18]. Antibody persistence upto 4 months after infection has been observed in studies from Iceland and USA. Later studies done using more sensitive CLIA test kits have shown antibody persistence for more than 6 months of SARS-CoV-2 infection.

This study was done using VITROS Ig G KIT in which results are calculated as signal/Cut off (S/Co). A serosurvey study on convalescent plasma therapy in COVID19 at Mayo clinic using the VITROS IgG kit revealed value between 1.0-4.64 S/Co as low titer, 4.62-18.45 as medium titer and above 18.45 as high titer [19]. In our study 40.6% came in low titer and 59.4% a medium titer.

## CONCLUSION

The seroprevalence of SARS-CoV-2 infection among HCWs in GMCT was found to be 19.1% in this study. Seropositivity was significantly higher in high risk group compared to low risk group. Antibody decay kinetics observed in this study is comparable to that in published literature. Even after six months of SARS-CoV-2 infection, 33.3% of patients had SARS-CoV-2 IgG antibody. The persistence of SARS-Cov-2 IgG in patients who were diagnosed to have COVID 19 at 5,4,3,2 and 1 month prior to the study period was 73%, 79.2%, 87%, 88.9% and 92.9% respectively. The high seropositivity among HCWs [19.1%] in GMCT compared to the average seropositivity among HCWs [10.5%] across the state reflects the infection prevention and control challenges in a hybrid hospital which caters to both COVID and non-COVID patients.

## Data Availability

All DATA has been archived

